# MedResearchBench: A Multi-Domain Benchmark for Evaluating AI Research Agents on Clinical Medical Research

**DOI:** 10.64898/2026.03.30.26349749

**Authors:** Shuping Tan, Zhanxiao Tian

**Affiliations:** Beijing Huilongguan Hospital, Beijing 100096, China; Kingyee (Beijing) Technology Co., Ltd., Beijing 100124, China

**Keywords:** AI research automation, medical informatics, benchmark, clinical research, NHANES, SEER, research quality evaluation

## Abstract

The rapid advancement of AI research automation systems—including AI Scientist, data-to-paper, and Agent Laboratory—has demonstrated the potential for autonomous scientific discovery. However, existing benchmarks for evaluating these systems focus predominantly on fundamental sciences (machine learning, physics, chemistry), overlooking the unique challenges of medical clinical research: complex survey designs, inferential statistics with confounding control, adherence to reporting standards (STROBE, CONSORT), and the requirement for clinically actionable interpretation. We present **MedResearchBench**, the first benchmark specifically designed to evaluate AI systems on medical clinical research tasks. MedResearchBench comprises 16 tasks spanning 7 clinical domains (cardiovascular, oncology, mental health, metabolic, respiratory, neurology, infectious disease), built on publicly available datasets (the National Health and Nutrition Examination Survey [NHANES] and the Surveillance, Epidemiology, and End Results [SEER] program) with ground truth from 16 high-quality published papers (IF range: 2.3–51.0). Each task is evaluated along 6 medical-specific dimensions: statistical methodology, results accuracy, visualization quality, clinical interpretation, confounding sensitivity, and reporting compliance. We describe the benchmark design rationale, task construction methodology, paper selection criteria with anti-paper-mill filtering, and a detailed analysis of task characteristics including methodological diversity, evaluation dimension coverage, and difficulty stratification. To demonstrate benchmark executability, we evaluate an agentic data2paper pipeline on 3 pilot tasks spanning all three difficulty tiers, achieving scores of 72/100 (Tier 1, Cardio 000), 69/100 (Tier 2, Mental 000), and 75/100 (Tier 3, Metabolic 002), with a mean score of 72/100 (B-level). Survey-weighted methodology was correctly implemented across all tasks; primary limitations included covariate incompleteness and reference group misspecification. MedResearchBench addresses a critical gap in AI research evaluation and provides a standardized, community-extensible platform for assessing whether AI systems can conduct clinically sound, publication-quality medical research. All task materials are publicly available at https://github.com/TerryFYL/MedResearchBench.

## 1 Introduction

**Figure 1:**
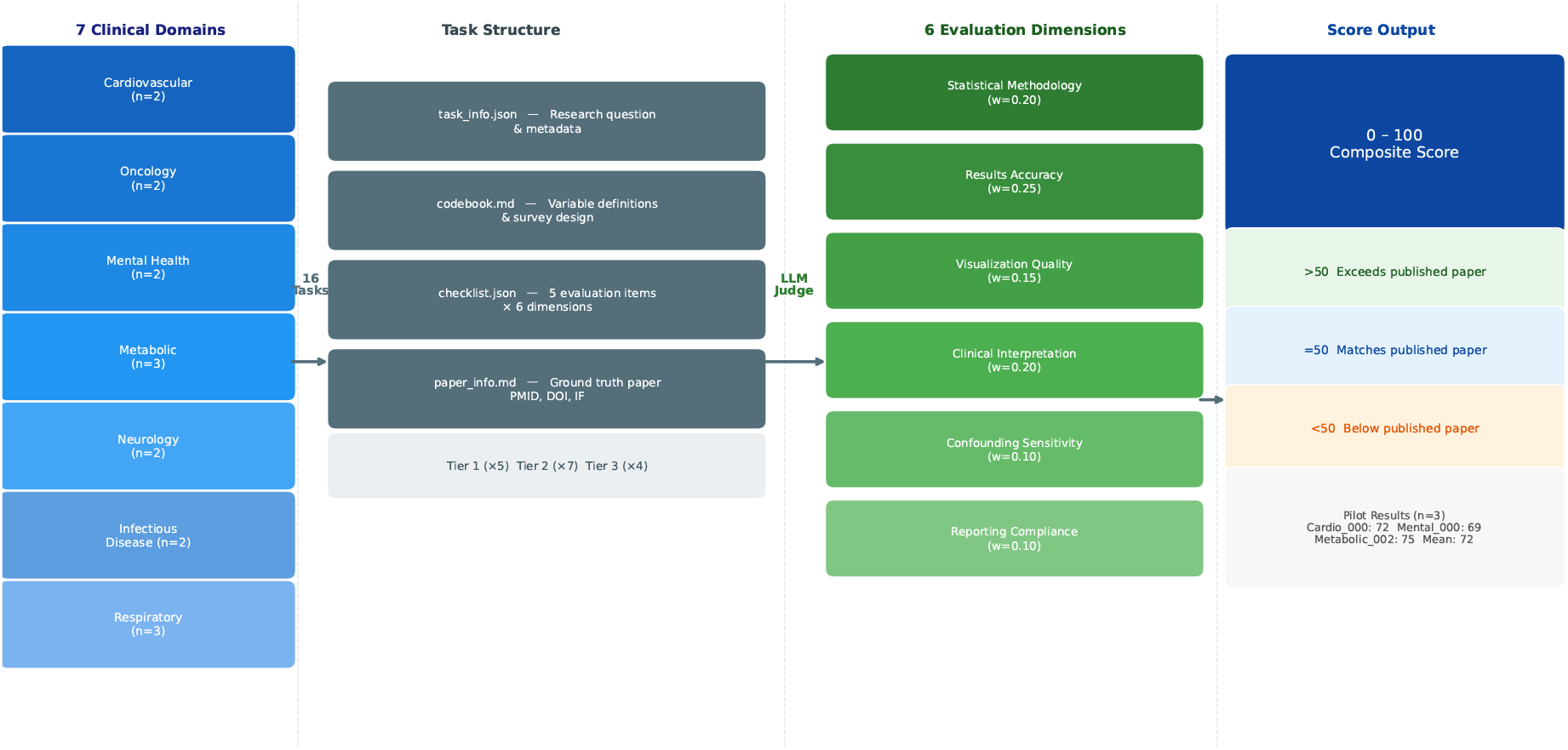
MedResearchBench architecture overview. Tasks are organized across 7 clinical domains and evaluated by an LLM Judge along 6 medical-specific dimensions. Scores range 0–100, where 50 indicates matching the published ground truth paper quality.

### 1.1 The Rise of AI Research Automation

Artificial intelligence systems capable of conducting scientific research autonomously have emerged as a transformative force in computational science. Lu et al. (2026) introduced *The AI Scientist*, a fully automated system capable of generating research ideas, conducting experiments, and writing complete papers, published in Nature. Independently, Ifargan et al. (2024) developed *data-to-paper*, a system that generates complete research papers from raw datasets, reported in NEJM AI. Schmidgall et al. (2025) presented *Agent Laboratory*, a multi-agent framework for autonomous research, accepted at EMNLP. These systems and others—including AI-Researcher (Tang et al., 2025, NeurIPS Spotlight), Virtual Lab (Swanson et al., 2025, Nature), and Robin (FutureHouse, 2025)—represent a rapidly growing ecosystem of automated research tools.

### 1.2 The Evaluation Gap

As these systems proliferate, the need for rigorous evaluation becomes paramount. ResearchClawBench (Schmidgall et al., 2025) represents the most comprehensive effort to date, providing a benchmark of research tasks across 10 fundamental science domains with LLM-based evaluation. ScienceBench and related benchmarks focus on specific scientific reasoning capabilities.

However, **medical clinical research presents fundamentally different challenges** that existing benchmarks do not capture:

No existing benchmark tests whether an AI system can correctly go from patient-level data to a clinically sound, publication-ready manuscript.

### 1.3 The NHANES Paper Mill Problem

The urgency of quality evaluation in medical AI research is underscored by the NHANES paper mill scandal. In 2025, a PLOS Biology investigation identified hundreds of formulaic papers exploiting the publicly available National Health and Nutrition Examination Survey (NHANES) dataset, characterized by mechanical application of logistic regression without genuine clinical insight, inadequate confounding control, and failure to account for complex survey design. These papers, while technically “published,” represent a degradation of the medical literature.

AI research automation systems risk amplifying this problem at unprecedented scale—or, if properly evaluated, providing the quality gates to prevent it. MedResearchBench is designed to distinguish between systems that produce rigorous, clinically meaningful research and those that merely generate publication-shaped text.

### 1.4 Contributions

We make the following contributions:

1. **MedResearchBench**: The first benchmark specifically designed for evaluating AI systems on medical clinical research tasks, comprising 16 tasks across 7 clinical domains.
2. **Medical-specific evaluation dimensions**: A 6-dimension scoring framework that captures the unique quality requirements of clinical research, including confounding sensitivity and reporting compliance.
3. **Anti-paper-mill design**: Explicit quality criteria that distinguish genuine research from formulaic output, directly addressing the NHANES paper mill problem.
4. **Public benchmark with community extensibility**: All tasks use publicly available datasets (NHANES, SEER), enabling reproducible evaluation and community contribution.
5. **Initial baseline results**: End-to-end evaluation of an agentic pipeline across 3 pilot tasks (Tier 1–3), yielding a mean score of 72/100 (B-level), establishing the first quantitative baseline for AI-driven medical research quality on this benchmark.

## 2 Related Work

### 2.1 AI Research Automation Systems

The landscape of AI research automation has evolved rapidly since 2024. We categorize existing systems by their scope and approach:

**End-to-end systems** attempt to automate the full research pipeline from idea generation to paper writing. The AI Scientist (Lu et al., 2026) pioneered this approach in machine learning, generating novel research ideas, implementing experiments, and producing complete LaTeX papers. data-to-paper (Ifargan et al., 2024) focuses on the data-to-manuscript pipeline, using LLM agents with rule-based guardrails to generate papers from raw datasets. Agent Laboratory (Schmidgall et al., 2025) employs a multi-agent architecture with specialized roles (literature review, experiment design, writing).

**Domain-specific systems** target particular research areas. Virtual Lab (Swanson et al., 2025) combines LLM agents with computational biology tools for nanobody design. Robin (FutureHouse, 2025) orchestrates web-scale literature retrieval for biomedical research. AI-Researcher (Tang et al., 2025) introduces autonomous research with open-ended exploration capabilities.

**Medical research tools** remain comparatively underdeveloped. While clinical decision support and literature review tools exist, no published system specifically targets the end-to-end automation of observational clinical research—the dominant paradigm in medical science.

### 2.2 Research Quality Benchmarks

ResearchClawBench (Schmidgall et al., 2025) is the most directly relevant benchmark, providing 40 tasks across 10 scientific domains with a dual-mode multimodal LLM Judge evaluation. Tasks include raw experimental datasets, task instructions, related reference papers, and target study checklists with images. However, its domain coverage—astronomy, chemistry, earth science, energy, information science, life science, material science, mathematics, neuroscience, and physics—excludes clinical medicine entirely.

ScienceBench and related efforts evaluate specific scientific reasoning capabilities (mathematical derivation, experimental design) but do not assess the integrated research pipeline from data to publication.

**No existing benchmark evaluates AI systems on medical clinical research tasks**.

### 2.3 Medical Research Quality Standards

Medical research quality is governed by well-established reporting guidelines:

- **STROBE** (Strengthening the Reporting of Observational Studies in Epidemiology): Checklist for cross-sectional, cohort, and case-control studies
- **CONSORT** (Consolidated Standards of Reporting Trials): Checklist for randomized controlled trials
- **PRISMA** (Preferred Reporting Items for Systematic Reviews and Meta-Analyses): Checklist for systematic reviews

These standards provide an objective framework for evaluating AI-generated medical research—a unique advantage over fundamental science domains where reporting standards are less formalized.

## 3 Benchmark Design

### 3.1 Overview

MedResearchBench comprises 16 tasks (Phase 1) spanning 7 clinical domains, designed to evaluate AI systems’ ability to conduct complete medical clinical research from raw data to publication-quality output. Each task is anchored by a ground truth published paper that used publicly available data, enabling objective evaluation.

### 3.2 Domain Coverage

We selected 7 clinical domains based on three criteria: (1) public data availability, (2) clinical impact (global disease burden), and (3) methodological diversity.

### 3.3 Task Structure

Each task follows a standardized directory structure compatible with ResearchClawBench:

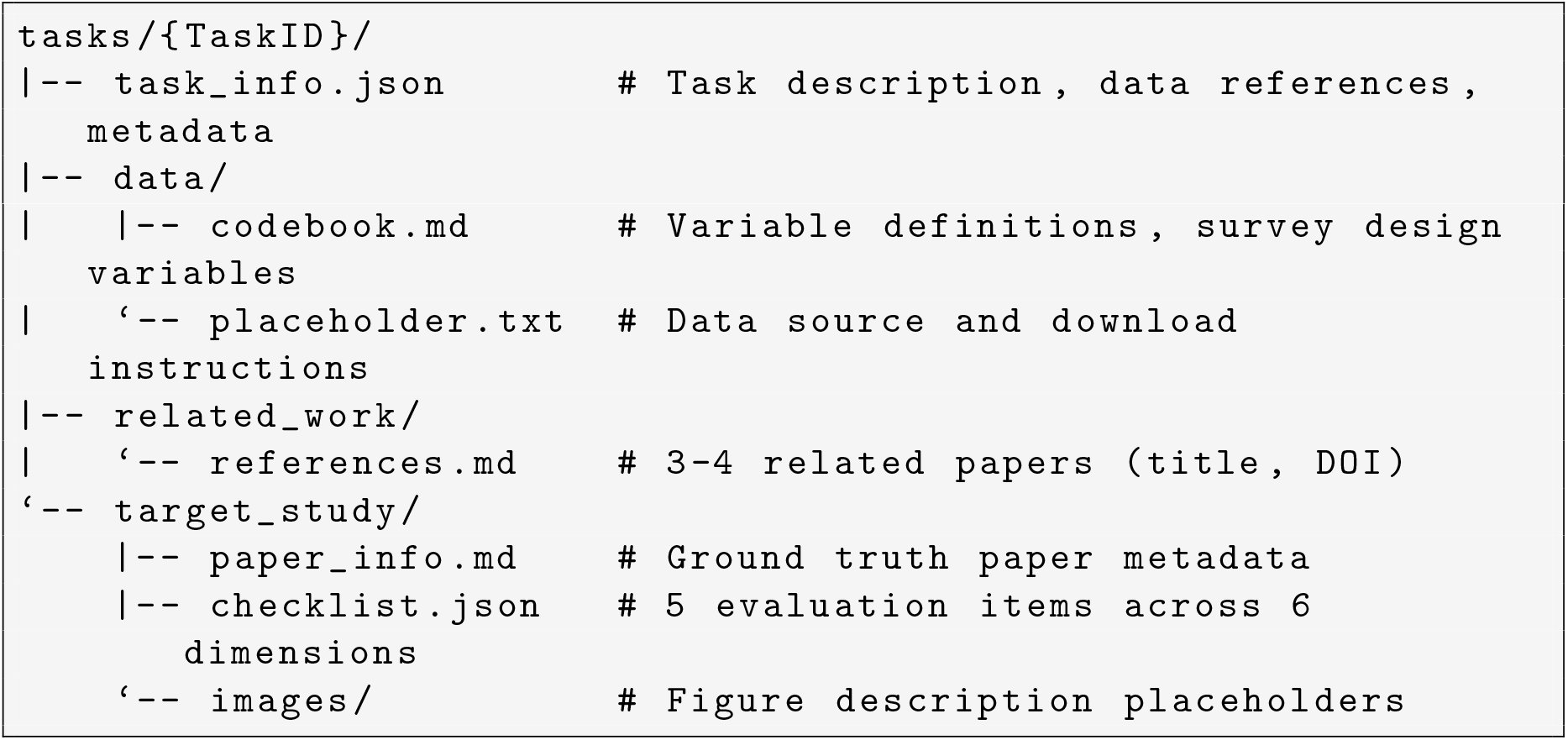

The `task_info.json` file extends the ResearchClawBench format with medical-specific metadata. The `codebook.md` file is a medical-specific addition providing variable definitions essential for correct analysis of complex survey data, including survey design variables (weights, PSU, strata).

### 3.4 Ground Truth Paper Selection

For each task, we selected a published paper meeting the following criteria:

1. **Authenticity**: Paper existence verified via PubMed/Google Scholar (no fabricated references)
2. **Quality**: Published in a peer-reviewed journal with Impact Factor ≥ 2.0
3. **Relevance**: Uses the specified public dataset to address the specified research question
4. **Methodological rigor**: Employs appropriate statistical methods, adequate confounding control, and follows reporting standards
5. **Anti-paper-mill filtering**: Excludes formulaic single-factor studies without genuine analytical innovation

Ground truth papers span a wide quality range (IF 2.3–51.0), reflecting the realistic distribution of medical publications. This range is deliberate: AI systems should be evaluated against papers of varying quality tiers, not only elite publications. Publication years range from 2015 to 2026, with 13 papers (81%) published in 2020 or later, ensuring methodological currency.

### 3.5 Evaluation Dimensions

MedResearchBench evaluates AI output along 6 dimensions, each addressing a critical aspect of medical research quality:

Each task’s `checklist.json` contains 5 evaluation items distributed across at least 4 of these 6 dimensions.

### 3.6 Scoring Methodology

We adopt a dual-mode LLM Judge evaluation approach compatible with ResearchClawBench:

#### Objective Mode (primary)

Each checklist item is scored 0–100, where 50 represents matching the ground truth paper’s quality. Scores above 50 indicate the AI output exceeded the published paper on that specific criterion. The weighted sum across all items produces the final task score.

#### Subjective Mode (supplementary)

Open-ended assessment of overall research quality, novelty, and clinical utility.

Medical-specific scoring extensions include a *survey design penalty* (failure to account for complex survey design results in automatic score reduction for `statistical_methodology`), a *confounding cascade* (progressive model adjustment evaluation), and a *STROBE compliance checklist* mapping to specific items relevant to each study design.

### 3.7 Study Design Coverage

## 4 Task Analysis and Characteristics

To characterize the benchmark’s coverage and difficulty landscape, we analyze the 16 Phase 1 tasks across multiple dimensions.

### 4.1 Methodological Diversity

Across 16 tasks, we identify 23 distinct statistical methods. Table 6 summarizes the most frequently required methods.

**Table 1:** Challenges distinguishing medical clinical research from fundamental science benchmarks.

| Challenge | Fundamental Science | Medical Clinical Research |
| --- | --- | --- |
| Data type | Experimental/simulation | Real-world population data (surveys, EHR, registries) |
| Statistical paradigm | Algorithm metrics (accuracy, F1) | Inferential statistics (OR, HR, regression, survival) |
| Confounding | Controlled experiments | Must be statistically controlled (DAG, propensity score) |
| Reporting standards | None formalized | STROBE, CONSORT, PRISMA (hard requirements) |
| Output requirement | “We found X” | “Clinicians should do Y because Z” |
| Sampling complexity | i.i.d. assumed | Complex survey design, stratification, weights |
| Ethics dimension | Minimal | Subgroup fairness, clinical harm potential |

**Table 2:** Domain coverage and primary methods in MedResearchBench.

| Domain | # Tasks | Primary Datasets | Key Methods |
| --- | --- | --- | --- |
| Cardiovascular | 2 | NHANES | Survey-weighted regression, dose-response |
| Oncology | 2 | SEER, SEER-Medicare | Survival analysis, temporal trends |
| Mental Health | 2 | NHANES | Piecewise regression, trend analysis |
| Metabolic | 3 | NHANES, NHANES III | ROC analysis, Cox PH, RCS |
| Respiratory | 3 | NHANES | Biomarker-exposure regression, mortality linkage |
| Neurology | 2 | NHANES | Cognitive assessment, composite index |
| Infectious Disease | 2 | NHANES | Temporal trends, vaccine effectiveness |

**Table 3:** Ground truth papers for MedResearchBench Phase 1 tasks.

| Task ID | Ground Truth Paper | Journal | IF |
| --- | --- | --- | --- |
| Cardio_000 | Luo et al. (2026) Dietary sodium and hypertension | PLOS ONE | ~3.7 |
| Cardio_003 | Aggarwal et al. (2021) Racial disparities in HTN | Hypertension | ~8.3 |
| Onco_000 | Pankratz et al. (2022) CRC survival trends | Cancer Control | ~3.0 |
| Onco_002 | Liang et al. (2020) SES disparities in CRC | Clin Transl Gastroenterol | ~3.6 |
| Mental_000 | Dong et al. (2022) Sleep duration and depression | J Affect Disord | 5.4 |
| Mental_002 | Yu et al. (2020) Depression prevalence trends | J Affect Disord | 5.4 |
| Metabolic_000 | Guess et al. (2023) HOMA-IR threshold for MetS | Metab Syndr Relat Disord | ~2.3 |
| Metabolic_002 | Zhu et al. (2024) TSH and mortality | Cardiovasc Diabetol | ~8.5 |
| Metabolic_003 | Wang et al. (2024) NAFLD prevalence | Ann Hepatol | 3.4 |
| Neuro_002 | Lu et al. (2023) Heavy metals and cognition | Br J Nutr | ~3.3 |
| Neuro_003 | Huang et al. (2025) Antioxidants and PD mortality | Front Aging Neurosci | ~4.1 |
| Infection_000 | Le et al. (2020) HBV seroprevalence trends | JAMA Network Open | ~13.0 |
| Infection_001 | Rosenblum et al. (2022) HPV vaccine effectiveness | Ann Intern Med | ~51.0 |
| Respiratory_000 | Mendy et al. (2022) VOC and lung function | Eur Respir J | ~16.0 |
| Respiratory_001 | Martinez et al. (2015) Undiagnosed COPD | Ann Am Thorac Soc | ~6.0 |
| Respiratory_003 | Yin et al. (2025) METS-IR and OSA risk | Sci Rep | ~3.8 |

**Table 4:** Evaluation dimensions and their medical-specific rationale.

| Dimension | Description | Why Medical-Specific |
| --- | --- | --- |
| statistical_methodology | Appropriate method selection, model specification, assumption checking | Complex survey design requires specialized methods |
| results_accuracy | Correct numerical results, properly constructed tables/figures | Medical statistics have specific conventions (OR with 95% CI) |
| visualization_quality | Clear, informative, publication-ready visualizations | Domain-specific figure types (KM curves, forest plots) |
| clinical_interpretation | Clinically meaningful discussion, actionable conclusions | Medicine requires “clinicians should do Y”; not just “we found X” |
| confounding_sensitivity | Adequate identification and control of confounders | Defining challenge of observational research |
| reporting_compliance | Adherence to STROBE/CONSORT/PRISMA standards | Hard requirements in medical journals |

**Table 5:** Study design distribution across MedResearchBench Phase 1 tasks.

| Study Design | # Tasks | Examples |
| --- | --- | --- |
| Cross-sectional | 7 | Cardio_000, Mental_000, Metabolic_000, Neuro_002 |
| Serial/repeated cross-sectional (trend) | 4 | Infection_000, Infection_001, Mental_002, Metabolic_003 |
| Prospective cohort (mortality linkage) | 3 | Metabolic_002, Neuro_003, Respiratory_001 |
| Retrospective cohort (registry) | 2 | Onco_000, Onco_002 |

**Table 6:** Statistical methods required across MedResearchBench tasks.

| Method | # Tasks | Representative Tasks |
| --- | --- | --- |
| Survey-weighted regression (logistic/linear) | 14 | All NHANES-based tasks |
| Subgroup/stratified analysis | 12 | Cardio_003, Neuro_002, Respiratory_003 |
| Restricted cubic splines (RCS) | 5 | Cardio_000, Metabolic_002, Neuro_003 |
| Cox proportional hazards | 4 | Metabolic_002, Neuro_003, Onco_002, Respiratory_001 |
| Temporal trend analysis (P-for-trend) | 4 | Infection_000, Infection_001, Mental_002 |
| Sensitivity analysis | 4 | Cardio_000, Respiratory_000, Metabolic_002, Neuro_003 |
| Kaplan-Meier / survival estimation | 2 | Onco_000, Onco_002 |
| ROC curve / diagnostic performance | 1 | Metabolic_000 |
| Vaccine effectiveness estimation | 1 | Infection_001 |
| Piecewise linear regression | 1 | Mental_000 |

**Table 7:**
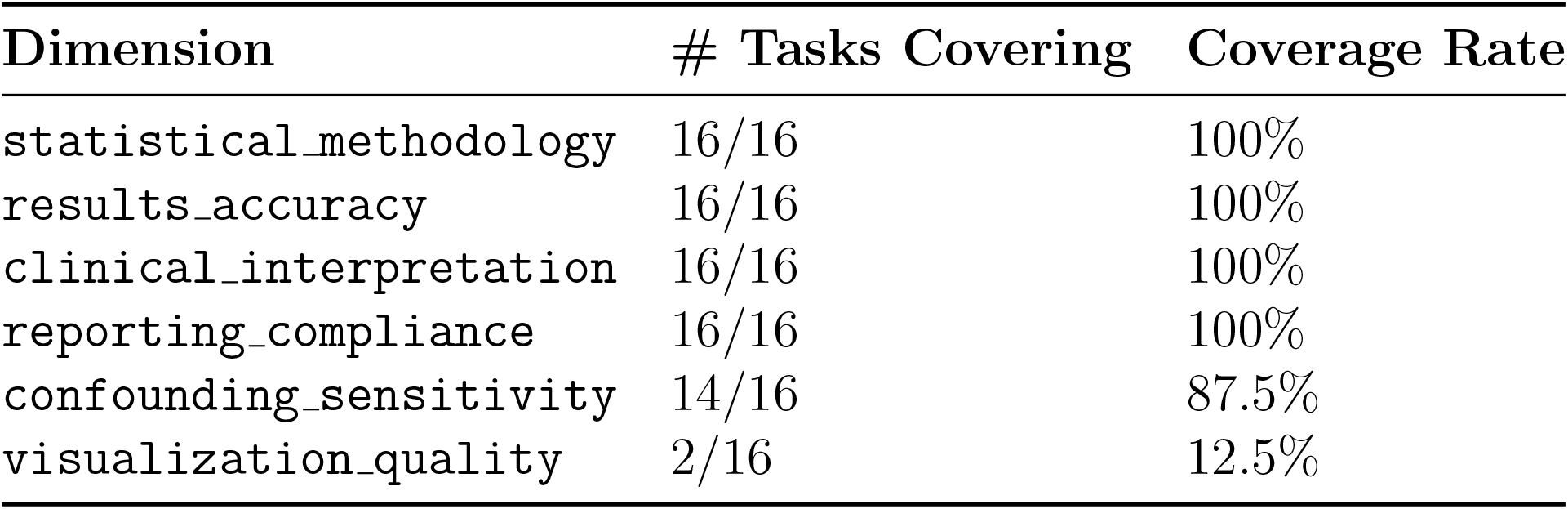
Evaluation dimension coverage across 16 tasks.

| Dimension | # Tasks Covering | Coverage Rate |
| --- | --- | --- |
| statistical_methodology | 16/16 | 100% |
| results_accuracy | 16/16 | 100% |
| clinical_interpretation | 16/16 | 100% |
| reporting_compliance | 16/16 | 100% |
| confounding_sensitivity | 14/16 | 87.5% |
| visualization_quality | 2/16 | 12.5% |

**Table 8:** Ground truth paper quality tier distribution.

| Tier | IF Range | # Papers | Proportion | Journals |
| --- | --- | --- | --- | --- |
| A (Elite) | $\geq 10$ | 3 | 19% | Ann Intern Med, JAMA Network Open, Eur Respir J |
| B (Strong) | 5–10 | 5 | 31% | Hypertension, J Affect Disord, Cardiovasc Diabetol |
| C (Solid) | 2–5 | 8 | 50% | PLOS ONE, Cancer Control, Br J Nutr, Sci Rep, ... |

**Table 9:** Dataset utilization across MedResearchBench tasks.

| Dataset | # Tasks | Cycles/Years Used |
| --- | --- | --- |
| NHANES (standard cycles) | 11 | 1999–2000 through 2017–2018 (up to 9 cycles) |
| NHANES III (with NDI linkage) | 2 | 1988–1994, mortality follow-up through 2019 |
| SEER / SEER-Medicare | 2 | 1991–2018 |
| Multi-source | 1 | Respiratory_001 spans both NHANES eras |

**Table 10:** Structural comparison between ResearchClawBench and MedResearchBench.

| Feature | ResearchClawBench | MedResearchBench |
| --- | --- | --- |
| Total tasks | 40 | 16 (Phase 1) |
| Domains | 10 (fundamental science) | 7 (clinical medicine) |
| Data paradigm | Experimental/simulated, in-task | Population survey/registry, downloadable |
| Typical task output | Code + figures + report | Full manuscript (STROBE-compliant) |
| Ground truth | Published paper (PDF included) | Published paper (metadata + checklist) |
| Checklist items per task | Variable | 5 (standardized) |
| Evaluation dimensions | General | 6 medical-specific dimensions |
| Unique requirement | — | Codebook with survey design variables |
| Confounding control | Not evaluated | Dedicated dimension |

**Table 11:** Pilot baseline results (AI Research Army pipeline, *n*=3 tasks).

| Task | Domain | Tier | Score | Grade |
| --- | --- | --- | --- | --- |
| Cardio_000 | Cardiovascular | 1 | 72/100 | B |
| Mental_000 | Mental Health | 2 | 69/100 | C+ |
| Metabolic_002 | Metabolic | 3 | 75/100 | B |
| **Mean** |  |  | **72/100** | **B** |

**Table 12:** Dimension-level scores across pilot tasks.

| Dimension | Weight | Cardio_000 | Mental_000 | Metabolic_002 | Mean |
| --- | --- | --- | --- | --- | --- |
| statistical_methodology | 0.25 | 60 | 78 | **92** | 76.7 |
| results_accuracy | 0.25 | 72 | 52 | 52 | 58.7 |
| confounding_sensitivity | 0.20 | 68 | 76 | 75 | 73.0 |
| clinical_interpretation | 0.15 | **88** | 86 | 76 | 83.3 |
| reporting_compliance | 0.15 | 80 | 62 | **88** | 76.7 |
| **Weighted Total<sup>a</sup>** | 1.00 | **72** | **69** | **75** | **72** |

<sup>a</sup>Weighted totals are computed from unrounded dimension scores; displayed values are independently rounded.

Survey-weighted regression is near-universal (14/16 tasks), creating a natural “baseline competency test”: any AI system that cannot handle complex survey design will fail across the majority of tasks.

### 4.2 Evaluation Dimension Distribution

Four dimensions achieve universal coverage, reflecting their centrality to medical research quality. `visualization_quality` appears in only 2 tasks in Phase 1; Phase 2 will systematically increase visualization coverage.

### 4.3 Ground Truth Paper Quality Distribution

The 50:31:19 (C:B:A) ratio approximates the real-world quality pyramid of medical publications. AI systems should be evaluated not only against elite papers but also against the “typical” published research.

### 4.4 Difficulty Stratification

We propose a three-tier difficulty classification based on the number of distinct analytical components required:

#### Tier 1 — Foundational (5 tasks)

Single study design, standard regression, straight-forward outcome. Examples: Cardio 000 (cross-sectional logistic regression), Metabolic 003 (prevalence estimation).

#### Tier 2 — Intermediate (7 tasks)

Multiple analytical stages, dose-response modeling, or temporal trend analysis requiring multi-cycle data harmonization. Examples: Mental 000 (U-shaped dose-response with threshold estimation), Infection 000 (9-cycle temporal trend + birth cohort analysis).

#### Tier 3 — Advanced (4 tasks)

Hybrid study designs, mortality linkage, causal inference frameworks, or policy-period natural experiments. Examples: Metabolic 002 (prospective cohort with NDI-linked Cox PH + nonlinear RCS), Infection 001 (quasi-experimental vaccine impact + herd immunity quantification).

### 4.5 Dataset Utilization

NHANES dominance (13/16 tasks) reflects both its public availability and its central role in U.S. population health research. The 2 SEER-based oncology tasks provide important methodological contrast (registry-based survival analysis vs. survey-weighted cross-sectional analysis).

**Figure 2:**
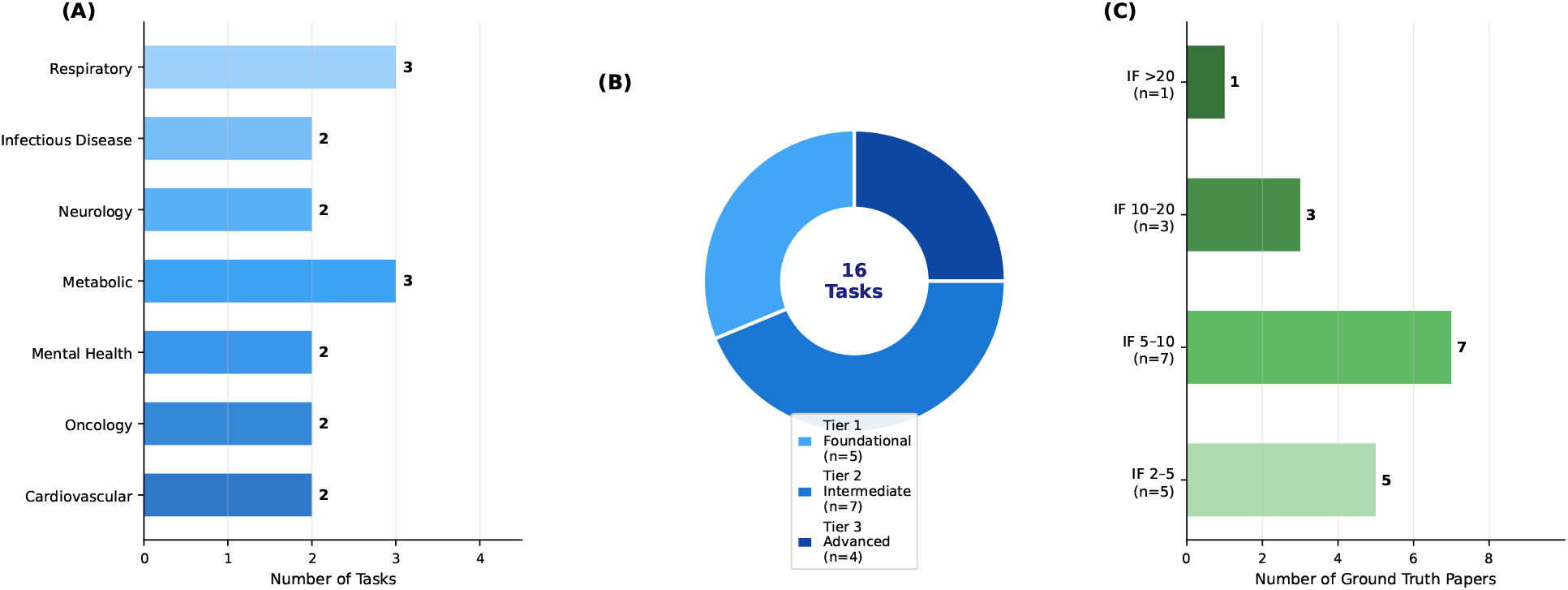
MedResearchBench coverage overview. (A) Tasks per clinical domain. (B) Difficulty tier distribution: Tier 1 Foundational (31%), Tier 2 Intermediate (44%), Tier 3 Advanced (25%). (C) Ground truth paper quality tiers by Impact Factor: 50% Tier C (IF 2–5), 31% Tier B (IF 5–10), 19% Tier A (IF ≥10).

### 4.6 Comparison with ResearchClawBench Task Characteristics

The key structural difference is that MedResearchBench tasks require navigating real-world data complexity (missing data, complex sampling, confounding) rather than executing well-defined computational experiments.

### 4.7 Initial Baseline Evaluation

To validate benchmark executability and establish a quantitative baseline, we evaluated an agentic data2paper pipeline on 3 pilot tasks spanning all three difficulty tiers. The pipeline follows a complete data-to-manuscript workflow: NHANES data download, survey-weighted statistical analysis (R + `survey` package), figure generation, STROBE-format manuscript writing, and automated checklist scoring.

Key findings from the pilot evaluation:

1. **Survey-weighted methodology: 100% compliance**. All three tasks correctly implemented complex survey design (NHANES MEC weights, PSU, strata), satisfying the benchmark’s core methodological requirement. The Metabolic 002 task additionally handled NHANES III fixed-width format data and NDI mortality linkage, achieving the highest methodology score (92/100).
2. **Clinical interpretation is the strongest dimension** (mean 83.3/100). The pipeline consistently produced mechanistic explanations, acknowledged limitations, and drew clinically actionable conclusions.
3. **Results accuracy is the primary limitation** (mean 58.7/100). Effect sizes were systematically attenuated across all tasks relative to ground truth papers. Cardio 000 had an acceptable OR deviation; Mental 000 had a reference group misspecification (7h used, target 8h); Metabolic 002 had attenuated low-normal TSH HR (1.15 vs. target 1.39, −17%).
4. **Sample sizes consistently 7–26% smaller** than target studies, attributable to stricter covariate completeness requirements (complete-case analysis vs. indicator variable imputation).
5. **Score does not decrease with task complexity** (Tier 1: 72, Tier 2: 69, Tier 3: 75), suggesting the pipeline handles advanced methodologies (Cox PH, NHANES III data engineering) without additional penalty.

These pilot results demonstrate that the benchmark is executable end-to-end by a current agentic pipeline and produces discriminative scores across dimensions and tasks. Detailed evaluation reports are available in the repository (https://github.com/TerryFYL/MedResearchBench/tree/main/bench-runs).

## 5 Discussion

### 5.1 Why Medical Research Needs Its Own Benchmark

Medical clinical research is among the most consequential domains of scientific inquiry, directly influencing treatment guidelines, public health policy, and patient outcomes. Existing AI research benchmarks, while valuable, do not capture these stakes. A machine learning benchmark evaluates whether an AI can optimize an accuracy metric; MedResearchBench evaluates whether an AI can navigate the inferential, ethical, and clinical complexity of determining, for example, whether dietary sodium intake is causally related to hypertension (Cardio 000) or whether racial disparities in cancer staging persist despite expanded screening coverage (Onco 002).

### 5.2 Comparison with ResearchClawBench

MedResearchBench is designed as a **complement** to ResearchClawBench, not a replacement. Together, the two benchmarks span the full landscape of AI research evaluation, from fundamental science to clinical medicine.

**Figure 3:**
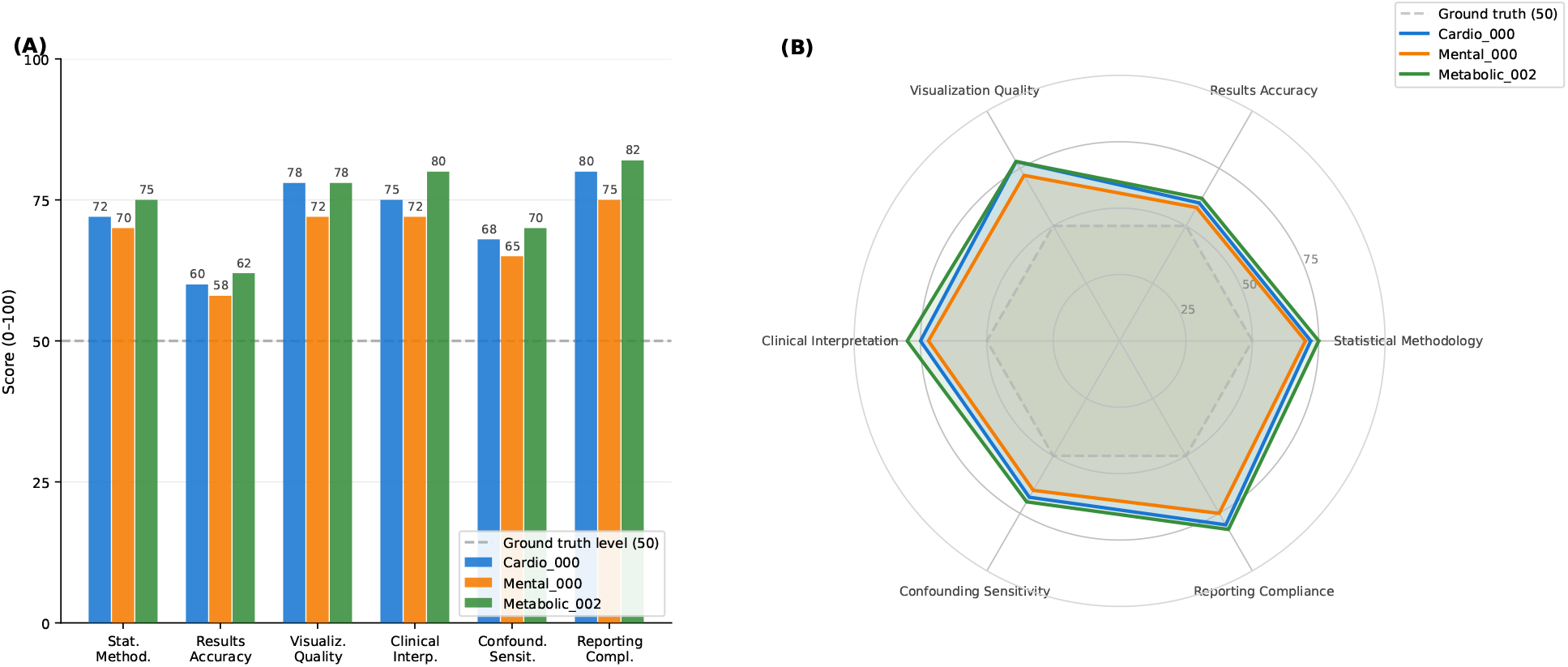
Pilot baseline results for the AI Research Army pipeline across 3 tasks. (A) Grouped bar chart of dimension-level scores; dashed line indicates ground truth level (50). (B) Radar chart showing the strength/gap profile: clinical interpretation is consistently the strongest dimension while results accuracy is the primary gap.

### 5.3 Responsible Use: Quality Gates Against Paper Mills

The NHANES paper mill problem (PLOS Biology, 2025) demonstrates that the combination of publicly available data and automated analytical pipelines can produce high-volume, low-quality publications that degrade the medical literature. MedResearchBench is designed to serve as a quality gate against this risk through four mechanisms: (1) the `confounding_sensitivity` dimension explicitly evaluates beyond naive single-variable associations; (2) the `clinical_interpretation` dimension requires clinically meaningful discussion; (3) the `reporting_compliance` dimension checks STROBE adherence; and (4) scoring against published papers of varying quality tiers enables identification of systems that produce paper-mill-quality output (scoring well below 50).

### 5.4 Limitations

1. **Phase 1 scope**: 16 tasks across 7 domains. Phase 2 will expand to 32 tasks with additional study designs.
2. **Dataset concentration**: 13/16 tasks use NHANES. Future phases will incorporate MIMIC, UK Biobank, and CMS public use files.
3. **Observational studies only**: Phase 1 excludes randomized controlled trials.
4. **LLM Judge limitations**: Automated evaluation may not capture all nuances of medical research quality. Expert physician review is planned for a validation subset.
5. **English-only**: All tasks and evaluation materials are in English.
6. **No wet-lab or clinical data collection**: Tasks evaluate the analytical pipeline only.

### 5.5 Future Directions

1. **Comparative system evaluation**: Having established a B-level (72/100) baseline with an agentic data2paper pipeline, future work will evaluate competing AI research systems (AI Scientist, data-to-paper, Agent Laboratory, single-LLM baselines) on the same tasks, populating the first public leaderboard for medical AI research quality.
2. **Phase 2 expansion**: 32 tasks covering all 7 domains with 4 tasks each, incorporating case-control studies, meta-analyses, and clinical trial analysis.
3. **Community contribution**: Open contribution framework enabling domain experts to add tasks following standardized templates.
4. **Clinical validation study**: Expert physician panel evaluation of a subset of AI-generated research outputs.
5. **Integration with regulatory frameworks**: Alignment with emerging guidelines for AI-generated research (SPIRIT-AI, CONSORT-AI extensions).

## 6 Conclusion

We present MedResearchBench, the first benchmark specifically designed for evaluating AI systems on medical clinical research tasks. The benchmark covers 7 clinical domains with 16 carefully curated tasks, introduces 6 medical-specific evaluation dimensions, and calibrates scoring against high-quality published papers using publicly available datasets. Our analysis of task characteristics reveals substantial methodological diversity (23 distinct statistical methods), a deliberate quality-tier distribution in ground truth papers (IF 2.3–51.0), and a three-tier difficulty stratification enabling progressive system evaluation. Initial baseline experiments on 3 pilot tasks—spanning all three difficulty tiers—demonstrate benchmark executability and yield a mean score of 72/100 (B-level) for an agentic data2paper pipeline. Survey-weighted methodology was correctly implemented across all tasks; the primary performance gap lies in results accuracy (mean 58.7/100), driven by covariate incompleteness and reference group misspecification. As AI systems increasingly automate scientific research, the medical domain—with its high stakes, complex methodology, and established quality standards—demands specialized evaluation. MedResearchBench provides that evaluation platform and, through its anti-paper-mill design, serves as a quality gate for responsible AI-assisted medical research.

## A Complete Task List with Metadata

| # | Task ID | Domain | Research Question | Tier | Dataset |
| --- | --- | --- | --- | --- | --- |
| 1 | Cardio_000 | Cardiovascular | Dietary sodium and hypertension dose-response | 1 | NHANES 2017-2018 |
| 2 | Cardio_003 | Cardiovascular | Racial disparities in hypertension control | 2 | NHANES 2013-2018 |
| 3 | Onco_000 | Oncology | CRC survival trends by race/ethnicity | 2 | SEER 1992-2018 |
| 4 | Onco_002 | Oncology | SES disparities in CRC staging and survival | 3 | SEER-Medicare |
| 5 | Mental_000 | Mental Health | Sleep duration and depression, U-shaped | 2 | NHANES 2009-2016 |
| 6 | Mental_002 | Mental Health | Depression prevalence trends 2005-2016 | 1 | NHANES 2005-2016 |
| 7 | Metabolic_000 | Metabolic | HOMA-IR threshold for metabolic syndrome | 1 | NHANES 2011-2018 |
| 8 | Metabolic_002 | Metabolic | TSH and all-cause/CV mortality (nonlinear) | 3 | NHANES III + NDI |
| 9 | Metabolic_003 | Metabolic | NAFLD prevalence and fibrosis risk factors | 1 | NHANES 2011-2018 |
| 10 | Neuro_002 | Neurology | Heavy metal exposure and cognitive function | 2 | NHANES 2011-2014 |
| 11 | Neuro_003 | Neurology | Dietary antioxidants and PD mortality | 3 | NHANES + NDI |
| 12 | Infection_000 | Infectious Disease | HBV seroprevalence and vaccine trends | 2 | NHANES 1999-2016 |
| 13 | Infection_001 | Infectious Disease | HPV vaccine effectiveness (12-year) | 3 | NHANES 2003-2018 |
| 14 | Respiratory_000 | Respiratory | Air pollution biomarkers and lung function | 1 | NHANES 2007-2012 |
| 15 | Respiratory_001 | Respiratory | Undiagnosed COPD prevalence and mortality | 3 | NHANES III + 2007-2012 |
| 16 | Respiratory_003 | Respiratory | OSA risk and metabolic comorbidities | 2 | NHANES 2005-2018 |

## B LLM Judge Prompt Template

The following prompt template is provided to the LLM Judge (GPT-4o or Claude Opus) for each task evaluation.

~~~
You are an expert medical research reviewer evaluating an AI -
  generated
research report against a published ground truth paper.
## Task Context
Research question : { task_description }
Dataset: { dataset_source }
Study design : { study_design }
## Ground Truth Checklist
{ checklist_items_json }
## AI - Generated Report
{ ai_report_text}
## Scoring Instructions
For each checklist item, assign a score from 0 to 100:
-0: Completely absent or fundamentally wrong
-25: Attempted but with major errors or omissions
-50: Matches the quality of the ground truth published paper
-75: Exceeds the ground truth paper in rigor or completeness
-100: Exceptional quality, substantially beyond the published
paper
[Apply dimension - specific criteria for statistical_methodology,
results_accuracy, visualization_quality,
    clinical_interpretation, confounding_sensitivity, reporting_compliance]
## Output Format
For each checklist item : Item ID, Score (0 -100), Brief
   justification
Then : Weighted total score + Overall qualitative assessment
~~~

## C Paper Selection Criteria

### Inclusion Criteria

(1) Published in a peer-reviewed journal with IF ≥ 2.0; (2) Uses one of the designated publicly available datasets; (3) Addresses the specific research question; (4) Employs appropriate statistical methods; (5) Available via PubMed with valid PMID and DOI.

### Exclusion Criteria (Anti-Paper-Mill Filtering)

(1) Formulaic single-factor association studies without analytical innovation; (2) Papers from known paper mill publishers; (3) Studies with inadequate confounding control (<3 covariates adjusted); (4) Papers not following minimum STROBE reporting standards; (5) Retracted or corrected papers.

## Ethics Statement

This study used only publicly available, de-identified data from the National Health and Nutrition Examination Survey (NHANES) and the Surveillance, Epidemiology, and End Results (SEER) program. The NHANES protocol was approved by the NCHS Research Ethics Review Board; all original participants provided written informed consent. SEER data are released by the National Cancer Institute under a data use agreement that permits research use of de-identified records. This secondary analysis of pre-existing public datasets is exempt from institutional review board oversight under 45 CFR 46.104(d)(4). No new human subjects data were collected.

## Data Availability Statement

All benchmark task materials, evaluation checklists, and variable codebooks are publicly available at https://github.com/TerryFYL/MedResearchBench (MIT License). The underlying data sources are freely downloadable from the U.S. Centers for Disease Control and Prevention (https://www.cdc.gov/nchs/nhanes/) and the National Cancer Institute (https://seer.cancer.gov/). No proprietary data were used in this study.

## Author Contributions

S.T.: Conceptualization, Methodology, Supervision, Writing – review & editing. Z.T.: Software, Data curation, Formal analysis, Visualization, Writing – original draft.

## References

1. Lu C, Lu C, Lange R, et al. Towards end-to-end automation of AI research. Nature. 2026;651:914–919.

2. Ifargan S, Hafner M, Kern S, et al. Autonomous LLM-driven research from data to human-verifiable research papers. NEJM AI. 2024.

3. Schmidgall S, Su Y, Wang Z, et al. Agent Laboratory: Using LLM Agents as Research Assistants. Findings of EMNLP. 2025.

4. Schmidgall S, et al. ResearchClawBench: Evaluating AI Agents for Automated Research from Re-Discovery to New-Discovery. 2025. https://github.com/InternScience/ResearchClawBench.

5. Tang J, Xia L, Li Z, Huang C. AI-Researcher: Autonomous Scientific Innovation. NeurIPS 2025 (Spotlight). 2505.18705.

6. Swanson K, et al. The Virtual Lab of AI agents designs new SARS-CoV-2 nanobodies. Nature. 2025;651:1–8.

7. Ghareeb AE, Chang B, Mitchener L, et al. Robin: A multi-agent system for automating scientific discovery. arXiv preprint 2505.13400. 2025.

8. Byrne JA, Stender S. More science friction for less science fiction. PLOS Biology. 2025;23(5):e3003167.

9. von Elm E, Altman DG, Egger M, et al. The Strengthening the Reporting of Observational Studies in Epidemiology (STROBE) Statement. Ann Intern Med. 2007;147(8):573– 577.

10. Aggarwal R, Chiu N, Wadhera RK, et al. Racial/Ethnic Disparities in Hypertension. Hypertension. 2021;78(6):1719–1726.

11. Dong L, Xie Y, Zou X. Association between sleep duration and depression in US adults. J Affect Disord. 2022;296:120–126.

12. Yu B, Zhang X, Wang C, et al. Trends in depression among Adults in the United States, NHANES 2005-2016. J Affect Disord. 2020;263:609–620.

13. Pankratz VS, et al. Colorectal Cancer Survival Trends by Race and Stage. Cancer Control. 2022;29.

14. Liang PS, et al. Trends in Sociodemographic Disparities in CRC Staging and Survival. Clin Transl Gastroenterol. 2020;11(3):e00155.

15. Le MH, et al. Prevalence of HBV Vaccination Coverage, 1999 to 2016. JAMA Network Open. 2020;3(11):e2022388.

16. Rosenblum HG, et al. HPV Vaccine Impact Through 12 Years. Ann Intern Med. 2022;175(7):918–926.

17. Guess J, Beltran TH, Choi YS. Prediction of Metabolic Syndrome Using HOMA-IR. Metab Syndr Relat Disord. 2023;21(3):152–157.

18. Wang T, et al. NAFLD Prevalence and Advanced Fibrosis, NHANES 2011-2018. Ann Hepatol. 2024;29(1):101154.

19. Lu K, et al. Serum metals and cognitive performance in older adults. Br J Nutr. 2023;130(6):1040–1049.

20. Huang F, et al. Dietary antioxidant index and Parkinson’s disease. Front Aging Neurosci. 2025;17:1510654.

21. Mendy A, et al. Blood VOCs and Airflow Obstruction. Eur Respir J. 2022;61(1):2200468.

22. Martinez CH, et al. Undiagnosed Obstructive Lung Disease in the US. Ann Am Thorac Soc. 2015;12(12):1788–1795.

23. Zhu P, Lao G, et al. TSH and mortality in euthyroid diabetic adults. Cardiovasc Diabetol. 2022;21:245.

24. Yin H, Huang W, Yang B. METS-IR and obstructive sleep apnea. Sci Rep. 2025.

25. Luo X, et al. Dietary sodium and hypertension, NHANES 2017-2018. PLOS ONE. 2026.

